# The impact of community asymptomatic rapid antigen testing on COVID-19 hospital admissions: a synthetic control study

**DOI:** 10.1101/2022.04.19.22274050

**Authors:** Xingna Zhang, Ben Barr, Mark Green, David Hughes, Matthew Ashton, Dimitrios Charalampopoulos, Marta García-Fiñana, Iain Buchan

**Affiliations:** Research Associate, Department of Public Health, Policy & Systems, University of Liverpool, Liverpool, UK; Professor in Applied Public Health Research, Department of Public Health, Policy & Systems, University of Liverpool, Liverpool, UK; Reader in Health Geography, Department of Geography & Planning, University of Liverpool, Liverpool, UK; Lecturer in Health Data Science, Department of Health Data Science, University of Liverpool, Liverpool, UK; Director of Public Health, Liverpool City Council, Liverpool, UK; Lecturer in Public Health, Department of Public Health, Policy & Systems, University of Liverpool, Liverpool, UK; Professor of Health Data Science, Department of Health Data Science, University of Liverpool, Liverpool, UK; Chair in Public Health and Clinical Informatics, Department of Public Health, Policy & Systems, University of Liverpool, Liverpool, UK

**Keywords:** COVID-19, SARS-CoV-2 rapid antigen testing, community testing, mass testing, hospital admissions, synthetic control

## Abstract

**Objective:** To analyse the impact on hospital admissions for COVID-19 of large-scale, voluntary, public open access rapid testing for SARS-CoV-2 antigen in Liverpool (UK) between 6^th^ November 2020 and 2^nd^ January 2021.

**Design:** Synthetic control analysis comparing hospital admissions for small areas in the intervention population to a group of control areas weighted to be similar in terms of prior COVID-19 hospital admission rates and socio-demographic factors.

**Intervention:** COVID-SMART (Systematic Meaningful Asymptomatic Repeated Testing), a national pilot of large-scale, voluntary rapid antigen testing for people without symptoms of COVID-19 living or working in the City of Liverpool, deployed with the assistance of the British Army from the 6^th^ November 2020 in an unvaccinated population. This pilot informed the UK roll-out of SARS-CoV-2 antigen rapid testing, and similar policies internationally.

**Main outcome measure:** Weekly COVID-19 hospital admissions for neighbourhoods in England.

**Results:** The intensive introduction of COVID-SMART community testing was associated with a 43% (95% confidence interval: 29% to 57%) reduction in COVID-19 hospital admissions in Liverpool compared to control areas for the initial period of intensive testing with military assistance in national lockdown from 6^th^ November to 3^rd^ December 2020. A 25% (11% to 35%) reduction was estimated across the overall intervention period (6^th^ November 2020 to 2^nd^ January 2021), involving fewer testing centres, before England’s national roll-out of community testing, after adjusting for regional differences in Tiers of COVID-19 restrictions from 3^rd^ December 2020 to 2^nd^ January 2021.

**Conclusions:** The world’s first voluntary, city-wide SARS-CoV-2 rapid antigen testing pilot in Liverpool substantially reduced COVID-19 hospital admissions. Large scale asymptomatic rapid testing for SARS-CoV-2 can help reduce transmission and prevent hospital admissions.

**Summary box:** *What is already known on this topic:*

– Previous studies on managing the spread of SARS-CoV-2 have identified asymptomatic transmission as significant challenges for controlling the pandemic.
– Along with non-pharmaceutical measures, many countries rolled out population-based asymptomatic testing programmes to further limit transmission.
– Evidence is required on whether large scale voluntary testing of communities for COVID-19 reduces severe disease, by breaking chains of transmission.

*What this study adds:*

– The findings of this study suggest that large scale rapid antigen testing of communities for SARS-CoV-2, within an agile local public health campaign, can reduce transmission and prevent hospital admissions.
– The results indicate that policy makers should integrate such testing into comprehensive, local public health programmes targeting high risk groups, supporting those required to isolate and adapting promptly to prevailing biological, behavioural and environmental circumstances.

## Introduction

Asymptomatic transmission of SARS-CoV-2 has been a significant challenge in managing the COVID-19 pandemic. Modelling studies, based on the original strain, had suggested that more than half of all transmissions in the community may arise from individuals without symptoms, whether pre-symptomatic or never symptomatic.[1] Non-pharmaceutical interventions (NPIs) intended to reduce the risk of transmission from people without symptoms, such as mask-wearing, social distancing, and restrictions on travel and access to public spaces and mass gatherings have therefore been necessary. However, there have been concerns over the potential harms to society and the economy from blunt strategies such as national lockdowns, including their effects on mental health and health inequalities.[2]

Among other NPIs, many countries have now implemented SARS-CoV-2 antigen rapid testing with lateral flow devices (LFDs) for people without symptoms to know if they are potentially infectious and to self-isolate,[3] therefore helping to reduce the spread of the virus.[4–7] There has been considerable scientific, public, and political debate over the mass use of LFDs – the potential harms from false negative and false positive results, sometimes confusing public health with clinical contexts, and the economic opportunity costs.[8–10] Most ‘mass testing’ debates and policies have, however, lacked controlled comparisons of key outcomes such as hospitalisation for tested versus untested populations sharing concurrent pandemic phases, with comparable contexts of the virus and immunity.

On 6^th^ November 2020, before populations were vaccinated, the UK Government piloted the first whole city voluntary community testing programme that was open to all residents and workers in Liverpool without symptoms of COVID-19.[8] The approach was bold, with an urgent need to generate evidence on (i) how popular ‘mass testing’ would be, (ii) whether small, controlled-environment studies of SARS-CoV-2 antigen LFD accuracy would be reflected in a real world public health setting, and (iii) if large scale asymptomatic testing would contain transmission and reduce adverse health outcomes. The early findings from this pilot informed the eventual national roll out of SARS-CoV-2 antigen rapid testing across the UK, as well as internationally.[11,12]

While popular overall, there were substantial inequalities in the uptake of this programme, with lower uptake among Black, Asian and Minority Ethnic (BAME) groups, deprived neighbourhoods, areas located further from test sites and areas containing populations less confident in using internet technologies.[9] Evidence from the pilot has showed that lateral flow tests were sufficiently accurate for the intended community testing purpose, although the number of missed cases with high viral load (despite being small) should be taken into account in high consequence settings.[10,13] The pilot also showed that the expected change in impact of false results with prevalence should be accommodated in agile, local testing policies.[14,15] Initial analyses of case rates indicated that community testing in Liverpool was associated with a reduction by around a fifth in cases up to the end of December 2020, and that this contrast with other parts of England disappeared as community testing rolled out nationally.[8,10] Case detection also increased by around a fifth over this period. Causal links between testing and transmission are difficult to make in the context of a complex intervention, especially as the pilot was accompanied by a major communication campaign that may have affected risk behaviours of those not testing as well as those using LFDs.

We are unaware of any existing research investigating whether large scale testing of communities for COVID-19, intended to reduce chains of transmission, reduces severe disease. In this study, we evaluate whether large-scale rapid testing for asymptomatic SARS-CoV-2 infection was effective at reducing hospital admissions.

## Methods

### Patient and Public Involvement

The implementation of COVID-SMART in Liverpool involved regular focus groups with residents run by Liverpool City Council in collaboration with University of Liverpool as detailed at https://www.liverpool.ac.uk/coronavirus/research-and-analysis/covid-smart-pilot/.

### Setting

COVID-SMART (Systematic Meaningful Asymptomatic Repeated Testing)[4] was introduced for all people living or working in the City of Liverpool, in the North West of England, from 6^th^ November 2020. Liverpool is the fourth most deprived out of 343 local authorities in England,[16] and at the time, the unvaccinated population had the highest COVID-19 case rate in the country. Introduction of SMART coincided with the start of the second national lockdown (5^th^ November until 2^nd^ December 2020).

### Data

Our primary outcome is the weekly number of hospital admissions for COVID-19 (International Classification of Diseases 10^th^ edition: ICD10 codes UO7.1 or UO7.2)[17] in England between 19^th^ November 2020 and 15^th^ January 2021 (intervention period plus two weeks to allow for average lead time from infection to hospitalisation), aggregated to Middle Layer Super Output Areas (MSOA), using Hospital Episode Statistics (HES) data provided by NHS Digital covering the period 4^th^ October 2020 to 17^th^ January 2021.[18]

In England, MSOAs are standard geographical units (with an average population of 7,200 people) nested within local authorities. We used hospital admissions as the primary outcome, including those with a primary diagnosis of COVID-19 having tested positive (UO7.1) or having been clinically diagnosed with COVID-19 (UO7.2).[19] This outcome is less affected by changes in levels of case detection than other outcomes, such as case rates, because the observation probability is less affected by factors such as choice/behaviour, testing capacity and testing practices. This is important as an objective of the intervention was to increase case detection.

In synthesising controls, we used data on six local area characteristics that could potentially influence uptake of testing, transmission, effectiveness of control measures, and vulnerability to hospitalisations. These included the English Indices of Multiple Deprivation (IMD) 2019 - a composite measure of socioeconomic disadvantage,[16] population density, the percentage of the population who were over 70 years old using 2019’s mid-year population estimates from the Office for National Statistics, the proportion of the population from BAME groups obtained from the 2011 Census, and the proportion of the population that had previously been admitted to hospital for a chronic disease (cardiovascular disease, diabetes, chronic kidney disease or chronic respiratory disease) between 2014-2018, using HES data as outlined above. To additionally account for potential differences in access to SARS-CoV-2 polymerase chain reaction (PCR) testing between areas prior to SMART, we used Local Authority data available from the UK government COVID-19 dashboard on the number of tests per capita in the seven weeks prior to the introduction of SMART.[20]

### Intervention

COVID-SMART was introduced after the UK Government selected Liverpool to pilot large-scale rapid testing of asymptomatic individuals for SARS-CoV-2 antigen. From 6^th^ November 2020, supervised self-testing with Innova SARS-CoV-2 rapid antigen lateral flow tests (LFT) was made available to everyone without symptoms living or working in the City of Liverpool. During the initial intensive testing period (6^th^ November to 3^rd^ December) the programme was deployed with the assistance of the British Army and was advertised across multiple media channels with communications also drawing attention to parallel PCR testing for people with symptoms. The initial plan to test 75% of the asymptomatic population in two weeks proved infeasible but the availability of testing was popular with the public. From 3^rd^ December 2020 the service was handed over to Liverpool City Council. The overall aim of the pilot was to reduce or contain transmission of the virus, with testing focusing on the following purposes: (1) ‘test-to-protect’ vulnerable people and settings (for example, people living in care homes); (2) ‘test-to-release’ contacts of confirmed infected people sooner from quarantine than the stipulated period (for example, key workers in quarantine); and (3) ‘test-to-enable’ careful return to restricted activities to improve public health, social fabric, and the economy (for example, mass gatherings).

Test-positive individuals were instructed to isolate for 10 days as per national guidance and to take a confirmatory PCR test. By 2^nd^ January 2021, 45% (n= 213,285) of the population had at least one LFT result registered, with 40% (n= 84,869) of people tested receiving more than one test. Over this period 5,110 cases (2.4% of all people tested) were identified as positive on LFT. Testing was particularly intensive up to 3^rd^ December when 25% of the self-declared asymptomatic population were tested for the first time in less than a month. The intervention was hypothesised to reduce hospital admissions by preventing onward transmission from the effective isolation of positive cases and their contacts, and from the associated publicity raising general awareness of COVID-19 risk behaviours and mitigations. A proportion of those cases prevented due to SMART would have resulted in admissions to hospital.

### Analysis

We apply the synthetic control method for microdata developed by Robbins et al. to estimate the effect of SMART on COVID-19 hospital admissions.[21,22] The synthetic control method is a generalisation of difference-in-difference methods, whereby an untreated version of the intervention areas (i.e. a synthetic control) is created using a weighted combination of areas that were not exposed to the intervention, and the intervention effect is estimated by comparing the trend in outcomes in the intervention areas to that in the synthetic control areas following the intervention.[23]

As there would be an expected time lag between the introduction of SMART and reduced hospital admissions, we assume the minimum plausible period from the start of the testing programme to the time when we might expect an impact on hospitalisation to be two weeks. We therefore compare the trend in admissions between the intervention and synthetic control areas after 19^th^ November 2020 (i.e. 14 days after SMART started on 6^th^ November). We estimated the intervention effect over two periods: (i) initial intensive testing period with military support (6^th^ November to 3^rd^ December 2020) and (ii) followed by civilian rollout involving fewer testing centres (6^th^ November 2020 to 2^nd^ January 2021). The initial testing period coincided with the national lockdown. We used the extended period to understand the extent to which impacts were sustained. From mid-December, asymptomatic community testing was gradually extended to other areas of the country. We therefore limited the follow up time to 2^nd^ January because after that time community testing in Liverpool was no longer being conducted at a higher rate than the rest of England, removing the intervention contrast with control areas (see Figure SF1 in Supplementary file for a comparison of LFT testing in Liverpool versus the rest of the country).

To construct the synthetic control group, we derive calibration weights to match the 61 MSOAs in Liverpool to areas outside Liverpool before the introduction of SMART. The weighting algorithm derives weights that meet two constraints. Firstly, the weighted average of each of the six local area characteristics, outlined above (IMD, population density, proportion of the population above 70 years, proportion from BAME groups, 5-year chronic condition hospital admission rate and PCR testing rate prior to intervention) in the control group is equal to the average for Liverpool.[21] Secondly, the total number of COVID-19 hospital admissions in the control group equals the number of hospital admissions in Liverpool for each of the seven weeks prior to 19^th^ November (preintervention period). Matching the synthetic control group on preintervention trends in hospital admissions for COVID-19 was important to minimise potential differences in unobserved characteristics.

From the pool of MSOAs used to construct the synthetic control, we excluded MSOAs within the Liverpool City Region (LCR) (139 MSOAs in five local authorities, other than Liverpool) to avoid spill-over effects of community testing on neighbouring areas. We also excluded MSOAs from the control group if they were within authorities with a mean LFT testing rate of more than 1 per 100 population per week between 6^th^ November and 2^nd^ January 2021 to minimise the potential impact of local asymptomatic pilot programmes elsewhere. This level of testing was at the 85^th^ percentile for all local authorities during this period with only 15% of local authorities testing above this level (see Figure SF2 in Supplementary File). 142 MSOAs were excluded in this way (2.2% of all non-intervention MSOAs). This left 6,290 MSOAs used in constructing the synthetic control group.

The Average Treatment Effect for the Treated (ATT) was then estimated as the difference in cumulative number of hospital admissions in the post intervention period in Liverpool, compared to the (weighted) cumulative number of admissions in the synthetic control group. To estimate the 95% confidence intervals and p-values we used permutation samples, by repeating the analysis through 250 placebo iterations randomly allocating MSOAs outside Liverpool to the intervention group, to estimate the sampling distribution of the treatment effect and calculating permuted p-values and confidence intervals.[22]

Following the end of the national lockdown on 2^nd^ December 2020, a three-tiered system of local restrictions was implemented. Liverpool entered less stringent Tier 2 (High alert) restrictions due to lower levels of COVID-19, whilst most similar areas entered Tier 3 (Very High alert) restrictions, which had a relatively large impact on transmission.[23] We therefore adjusted our analysis for the extended period (6th November 2020 to 2nd January 2021) to remove the effect of the Tier 3 restrictions relative to Tier 2 restrictions, in the synthetic control group. Extending our previous analysis,[23] we find that Tier 3 restrictions reduced hospital admission rates by 17% (95% CI 13% to 21%) relative to Tier 2 restrictions, and that these effects started around the 17^th^ December 2020 and extended to the 21^st^ February 2021. We therefore adjusted the cases in Tier 3 areas upwards by this percentage during this period before deriving weights as outlined above to provide a synthetic control group reflecting transmission conditions that were experienced in Liverpool at that time (details for estimating this adjustment are given in Supplementary File, part 2).

All analysis was performed using R version 4.0.3 and the Microsynth package.[21]

### Sensitivity analyses

In sensitivity analysis we repeated the synthetic control models for the upper and lower plausible estimates of the potential effect of less stringent Tier 2 restrictions in Liverpool. These were based on the upper (21%) and lower (13%) bounds of the 95% confidence interval of our estimate of the tiered effect (results are provided in Supplementary File, part 4). We also replicated the analysis without excluding places with mean LFT rates above 1 per 100 population per week (results are provided in Supplementary File, part 5).

## Results

Table 1 presents summary statistics for the 61 MSOAs that make up Liverpool and the pool of MSOAs in the rest of England from which the synthetic control group was constructed. Liverpool has markedly higher levels of deprivation, higher population density, higher proportion of the population who had previously been admitted for a chronic disease, lower proportion of the population over 70 and a lower proportion of the population from BAME groups. In the seven weeks before the introduction of SMART, Liverpool had a higher number of PCR tests per capita, higher COVID-19 admission and case rates than the average for the rest of England. Because of the matching algorithm used to construct the synthetic controls, the weighted average of each variable in Table 1 achieved an exact match between the intervention (Liverpool) and synthetic control areas. Figure SF4 in Supplementary file shows the geographical pattern of these weights in constructing the synthetic control group.

**Table 1.** Comparison between Liverpool and the MSOAs in the rest of England used to construct the synthetic control group (i.e. excluding those within Liverpool City Region or with a high LFT testing rate).

|  | **Liverpool** | **MSOAs in the rest of England used to construct the synthetic control** |
| --- | --- | --- |
| **Number of MSOAs** | 61 | 6,290 |
| **Total population** | 498,042 | 52,330,147 |
| **2019 IMD score** | 43 | 21 |
| **Population density<sup>1</sup>**<br>**(people per hectare)** | 55 | 36 |
| **% of population 70+<sup>1</sup>** | 11 | 14 |
| **% BAME<sup>2</sup>** | 11 | 14 |
| **% of population with at least 1 admission for a chronic disease<sup>3</sup>** | 24 | 20 |
| **Number of PCR tests per 100,000 population<sup>4</sup>** | 3,572 | 2,552 |
| **Average hospital admissions per 100,000 population per week for COVID-19<sup>4</sup>** | 26 | 9 |
| **Weekly COVID-19 cases per 100,000 population per week<sup>4</sup>** | 464 | 203 |

<sup>1</sup> calculated using 2019's mid-year population estimates from the Office for National Statistics
<sup>2</sup> data obtained from the 2011 Census
<sup>3</sup> based on HES data between 2014 and 2018
<sup>4</sup> data refer to the pre-intervention period from 4<sup>th</sup> October 2020 to 5<sup>th</sup> November 2020

Figure 1 shows the trend for the average COVID-19 hospital admission rates from 5^th^ October 2021 until 15^th^ January across MSOAs in Liverpool, and the synthetic control group. Due to an exact match in calibration weights, trends were identical in the synthetic control and intervention group in the pre-intervention period (5^th^ October to 6^th^ November). However, trends began to diverge two weeks after the introduction of SMART with hospitalisations being lower in Liverpool than in the synthetic control. The lower trend in Liverpool continued throughout December before sharply rising in January to match the synthetic control as community testing expanded to other areas.

**Figure 1.**
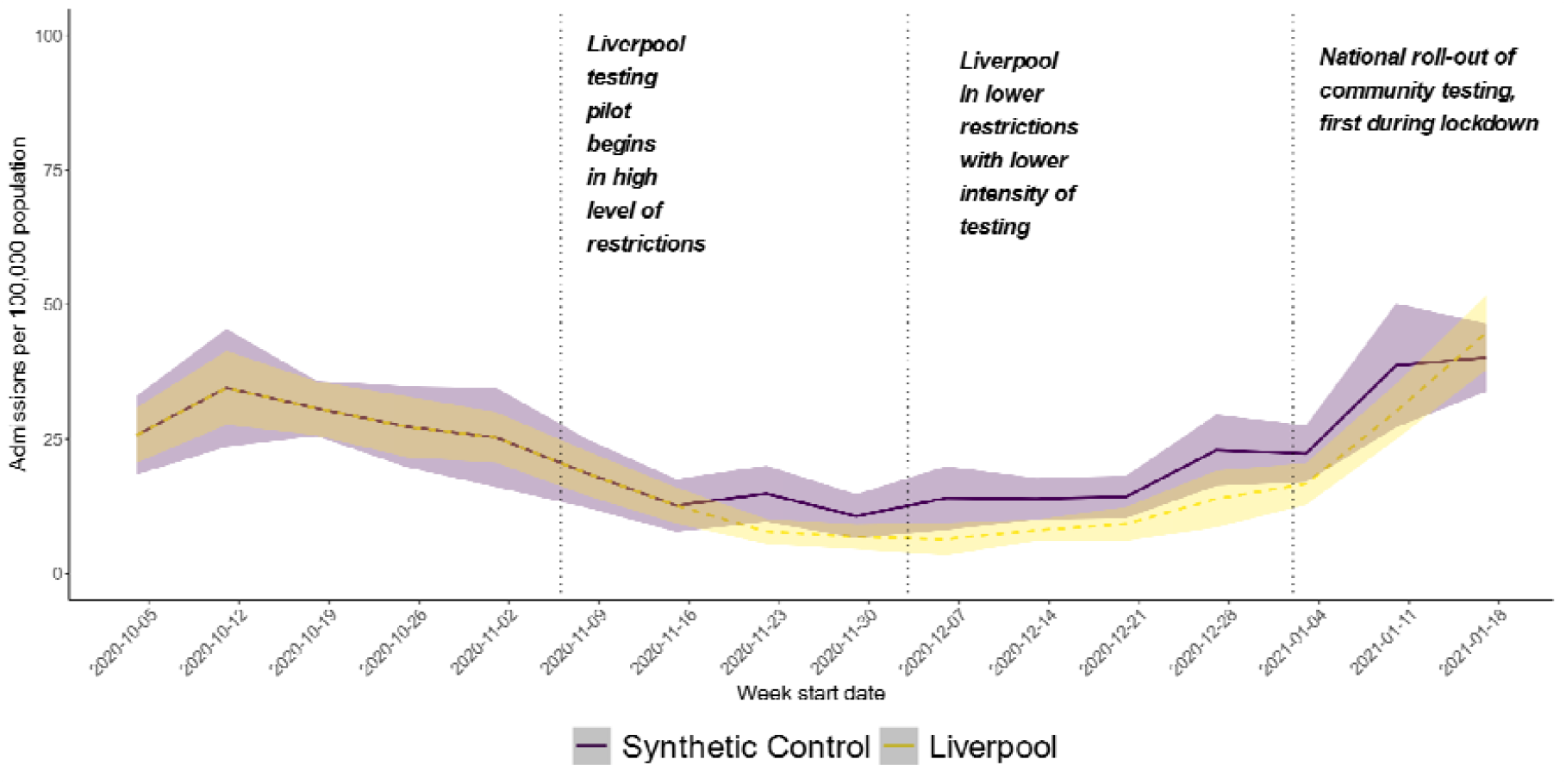
Trend in weekly COVID-19 hospital admission rates in MSOAs in Liverpool City compared to a synthetic control group constructed from the weighted average of MSOAs outside Liverpool City Region without community testing. Dotted vertical lines represent start of Liverpool community testing pilot on 6th November 2020, followed by Tier 2 restrictions on 3rd December 2020, before the national roll-out of community testing in lockdown on 3rd January 2021.

Table 2 shows the results of the synthetic control analysis, indicating estimated effect of SMART on COVID-19 hospital admissions.

**Table 2.** Estimated effects of COVID-SMART community testing on COVID-19 hospital admissions from synthetic control analysis, under alternative assumptions over the effects of a lower level of restrictions in Liverpool City in December 2020

| **Model** | **Intervention period** | **Assumed reduction in COVID-19 hospital admissions from Tier 3 vs 2 restrictions** | **% difference in COVID-19 hospital admissions between Liverpool and control** | **Lower 95% CL** | **Upper 95% CL** | **P- value** |
| --- | --- | --- | --- | --- | --- | --- |
| **1** | 6<sup>th</sup> November to 3<sup>rd</sup> December 2020 | Nil | - 43 % | - 57% | - 29% | <0.001 |
| **2** | 6<sup>th</sup> November 2020 to 2<sup>nd</sup> January 2021 | Nil | - 16 % | - 27% | 0% | 0.068 |
| **3** |  | 17% (central estimate) | - 25 % | - 35 % | - 11 % | <0.001 |

Over the initial intensive testing period (6^th^ November to 3^rd^ December 2021) (model 1), admission rates in Liverpool were 43% lower compared to the synthetic control group (95% CI 57% lower to 29% lower). In absolute numbers this 43% reduction is the equivalent of 146 fewer admissions in the period up to 3^rd^ December 2020.

When extending the analysis to the period up to January 2^nd^ (model 2), we observed a smaller, estimated effect of SMART reducing admissions by 16% (95% CI 27% lower to 0%) in Liverpool compared with control areas. After adjusting for the anticipated effect of Tier 3 restrictions on COVID-19 hospitalisations using our central estimate, the impact of community testing increased, reducing hospital admissions by 25% (95% CI 35% lower to 11% lower) (model 3).

Sensitivity analyses using the upper and lower plausible estimates of the potential Tier effect showed coherent and similar results with that of model 3 (see more details in Supplementary File, part 4). We also repeated our analysis by including areas with mean weekly LFT rates above 1 in 100 population; findings were similar to those presented above (see more details in Supplementary File, part 5).

## Discussion

We found that the introduction of community testing in Liverpool, ahead of its wider implementation across the UK, was associated with a reduction in hospital admissions for COVID-19 compared to what would have been expected in the absence of this intervention. This effect was greater when analysis was restricted to the first month of implementation, when testing was more intensive through military assistance and before Liverpool entered a lower level of restrictions than most other cities, at the same time as the Alpha variant spread nationally. This suggests that widespread community testing has an effect at reducing transmission and consequently admissions to hospital for COVID-19. And we found similar effects when we explored the impact of the early roll-out of community testing across the wider Liverpool City Region using an equivalent synthetic control analysis, where we estimated a 32% (95% CI: 22% to 39%) reduction in COVID-19 hospital admissions.[8] Early findings from this pilot informed the national roll-out of SARS-CoV-2 antigen rapid testing across the UK, and have influenced policies internationally.[11]

Our analysis has several strengths. We were able to use small area data to construct a control group that had very similar characteristics to our intervention population. The synthetic control approach ensured that control areas were similar in terms of both level and prior trends in hospital admissions, indicating that these areas were likely to have been affected by similar SARS-CoV-2 transmission patterns prior to the introduction of SMART in Liverpool. This is important as the parallel trends assumptions of simple difference-in-difference methods are not sufficient for analysis of infectious diseases, where the rate of change is intrinsically linked to the levels of infection at baseline.[24]

There are several limitations. Firstly, although we were able to match areas to ensure a good balance of potential confounding factors prior to the intervention, it is possible that concurrent changes in the intervention and/or control populations could bias the results. The major policy change that affected transmission at this time was the introduction of tiered restrictions and we have sought to adjust for these in our analysis and present sensitivity analysis assuming different effects of this policy on transmission. The adjustments we make for these differences in restrictions assume the effect of Tier 2 restrictions on transmission in Liverpool was the same as the average effect across Tier 2 areas in England. The effect could have, however, been greater in Liverpool, because unlike other Tier 2 areas, most of the areas surrounding Liverpool were in Tier 3. Being an island of lower restrictions may have seen surrounding populations using the restaurants and other facilities open in Liverpool that were closed in their own areas at the time. In addition, some other areas saw Tier 4 restriction in late December 2020. So, our estimates for the effect of SMART may be overly conservative.

Secondly, there are potential spill-over effects with community testing affecting transmission beyond Liverpool, particularly as it was available to people working in Liverpool. We have sought to account for that by excluding surrounding areas from the control group.

Thirdly, our synthetic control group was made up of a non-adjacent set of neighbourhoods, that in aggregate had followed similar trends in hospitalisation to the contiguous neighbourhoods of Liverpool. Our analysis did not consider potential effects on transmission of these differences in the spatial dispersion of the intervention and control neighbourhoods. One might expect that this would lead to a more rapid increase in transmission in Liverpool compared to the synthetic control areas, as neighbourhoods in Liverpool tended to be adjacent to areas with high case rates, whilst the synthetic control neighbourhoods tended to be adjacent to areas with lower case rates. In other words, this would be expected to dilute the intervention effect.

Fourthly, we were only able to use data on small neighbourhood areas, rather than on individuals and therefore were unable to investigate how effects of community testing varied by individual or household characteristics.

Finally, the causal inference cannot be tied down to the use of rapid antigen tests alone because the extensive communication required to implement community testing may have affected COVID-19 risk behaviours in those not taking tests.

There has been widespread debate about the potential benefits and harms of ‘mass testing’ with lateral flow tests in COVID-19 responses.[25] It is increasingly recognised that this can be an important non-pharmaceutical intervention for identifying infectious cases.[4,26–28] Given the importance of asymptomatic transmission,[1] measures that shorten the time between testing and results, have the potential to break chains of transmission through the timely isolation of the most infectious cases and their close contacts. Criticism of this approach, however, has focused on the accuracy of the tests, potential lack of adherence to self-isolation of those identified and their contacts and insufficient evaluation prior to roll-out.[5] The experience in Liverpool indicates that widespread community testing is feasible and leads to the detection of cases who would not otherwise have been identified.[8] Survey results from Liverpool indicated that a high proportion of cases identified reported that they did self-isolate after testing positive.[8]

It is plausible that the main effect in our analysis is causally related to the SMART intervention, especially as the study period pre-dates the main COVID vaccination roll-out. Over the full follow up period a 25% reduction of what would have been without SMART is the equivalent to an absolute reduction of approximately 239 admissions in Liverpool. Assuming an Infection Hospitalisation Ratio of 3.5%,[29] a reduction in 239 admissions would suggest that around 6,829 infections would need to be prevented to reduce hospital admissions by this amount. In other words, if this effect was causal, the isolation of the 5,691 positive cases identified through SMART is equivalent to preventing around 5,691 to 6,829 infections (up to 20% higher, assuming that Liverpool, a city in North West England, had an average reproduction rate of 1.0 to 1.2 during this study period)[30] overall due to breaking modes of transmission through catching cases earlier.

Although large-scale rapid testing for SARS-CoV-2 antigen has been implemented in many countries, there has been limited evaluation of its impact on transmission or hospitalisation. A modelling study found that one round of mass testing may reduce daily infections by up to 20–30%, but that these effects are likely to be relatively short lived with infections returning to pre mass-testing levels shortly after an initial wave of testing.[6] Analysis of population wide testing in Slovakia indicated that it was associated with a 58% reduction in transmission, although this analysis was not able to distinguish between the impact of mass testing and other control measures that were introduced at the same time.[7] Our study findings are broadly consistent with these estimates.[14]

Similar to previous studies we find that effects seem to have been greatest early on in the programme when large numbers of tests were administered to a large number of people within a relatively short time period.[6,31] Others have also highlighted the importance of combining rapidly testing in a large proportion of the population along with minimising test delays to control transmission, with modelling indicating that this can compensate for lower test sensitivity.[31] Clearly the use of asymptomatic testing to control transmission will be undermined if the positive cases identified are unable to or disinclined to isolate, for example if there are financial penalties to isolation. Effectiveness will also be reduced when, as we found in Liverpool, uptake of testing tends to be lower in populations where transmission tends to be higher (for example amongst more deprived groups).[9] Our findings suggest, however, that even where uptake is unequal and there are barriers to effective isolation, widespread community testing can potentially reduce transmission and subsequent hospitalisations at least in the short term. Further strategies for community asymptomatic testing should aim to maintain high levels of repeat testing, particularly targeted at high-risk groups. Combining this with other control measures could allow us to maintain control of SARS-CoV-2, whilst maintaining social and economic activity.

## Conclusion

The world’s first voluntary, city-wide SARS-CoV-2 rapid antigen testing pilot in Liverpool substantially reduced COVID-19 hospitalisation. Community asymptomatic testing for SARS-CoV-2 antigen with lateral flow devices has been a useful addition to the measures for mitigating COVID-19 risks. The success of such control measures relies on high levels of uptake and effective support to enable isolation of infectious people and their close contacts. For effective public health responses to COVID-19, testing is more than a test. It is a complex intervention comprising communication, technology and social responses.

## Data Availability

All the data we used are publicly accessible from https://coronavirus.data.gov.uk/ and statistical code is available from the corresponding author upon request.

## Declarations

### Transparency statement

BB and IB affirms that the manuscript is an honest, accurate, and transparent account of the study being reported; that no important aspects of the study have been omitted; and that any discrepancies from the study as originally planned have been explained.

### Competing interests

The City of Liverpool received a donation from Innova Medical Group towards the foundation of the Pandemic Institute. None of this funding has supported the research reported here or any of the authors’ work to date.

### Ethics statements

This study only used anonymised and aggregate data and did not require ethical approval.

### Patient consent for publication

Not required.

### Funding

This report is independent research funded by the Department of Health and Social Care. This work was supported by the Economic and Social Research Council [grant number ES/L011840/1]. IB is supported by the National Institute for Health Research as Senior Investigator. BB, XZ are supported by the NIHR Applied Research Collaboration North West Coast (ARC NWC). IB, BB and XZ are also supported by the National Institute for Health Research (NIHR) Health Protection Research Unit in Gastrointestinal Infections, a partnership between Public Health England, the University of Liverpool and the University of Warwick (Grant No: NIHR ref NIHR200910). The NIHR had no role in the study design, data collection and analysis, decision to publish or preparation of the article. The views expressed in this publication are those of the author(s) and not necessarily those of the NHS, NIHR or the Department of Health and Social Care.

### Authors’ contributions

All co-authors have fulfilled authorship criteria per ICMJE guidelines, have read and approved the final manuscript. BB conceptualised the analytic plan, analysed the data, interpreted the results, drafted the paper, and critically reviewed updated versions of the paper. IB had the general supervision of the study, provided expertise on clinical interpretation of findings, drafted and critically reviewed the paper. BB and IB are guarantors of the study. XZ cleaned and analysed the data, drafted and revised the paper. DC drafted the paper and helped with interpretation of findings. MGF provided expertise on study methodology and critically reviewed the paper. MG, and DH contributed to the analyses, helped with interpretation of findings and critically reviewed the paper.

### Data sharing

All the data we used are publicly accessible and statistical code is available from the corresponding author upon request.

## Acknowledgements

We thank all the Liverpool residents who participated during the study period. We also thank Liverpool City Council officials and Prof Matthew Ashton, Director of Public Health in Liverpool City for the support in the planning and delivery of the COVID-SMART programme.

## Supplement

### Part 1: Lateral flow testing rates over time

Trends in the number of tests over time (Figure SF1) reflect initial high uptake during the initial push, declining following planned withdrawal of military assistance shortly after Liverpool’s move into less stringent (Tier 2) local restrictions (announced 26^th^ November 2020, enacted 2^nd^ December 2020). Uptake remained initially low in December, before a sharp increase in the week before Christmas as individuals may have sought tests before mixing among Christmas bubbles. High demand was sustained after Christmas and into the national lockdown (starting 6^th^ January 2021).

**Figure SF1.**
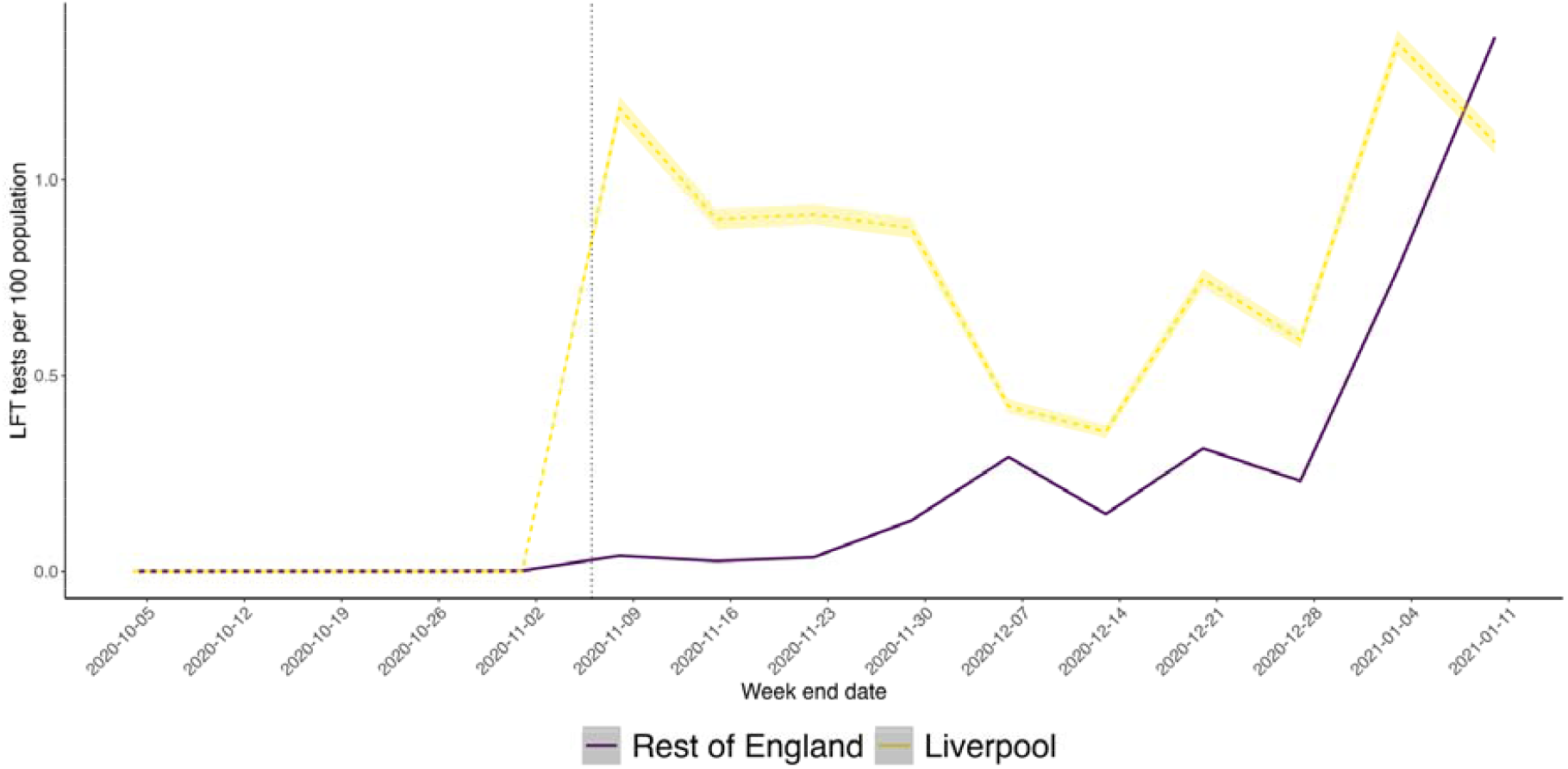
Trend in lateral flow tests per 100 population each week in Liverpool and in the rest of England. The vertical line marks the rollout of the asymptotic community testing pilot in Liverpool on 6^th^ November 2020.

**Figure SF2.**
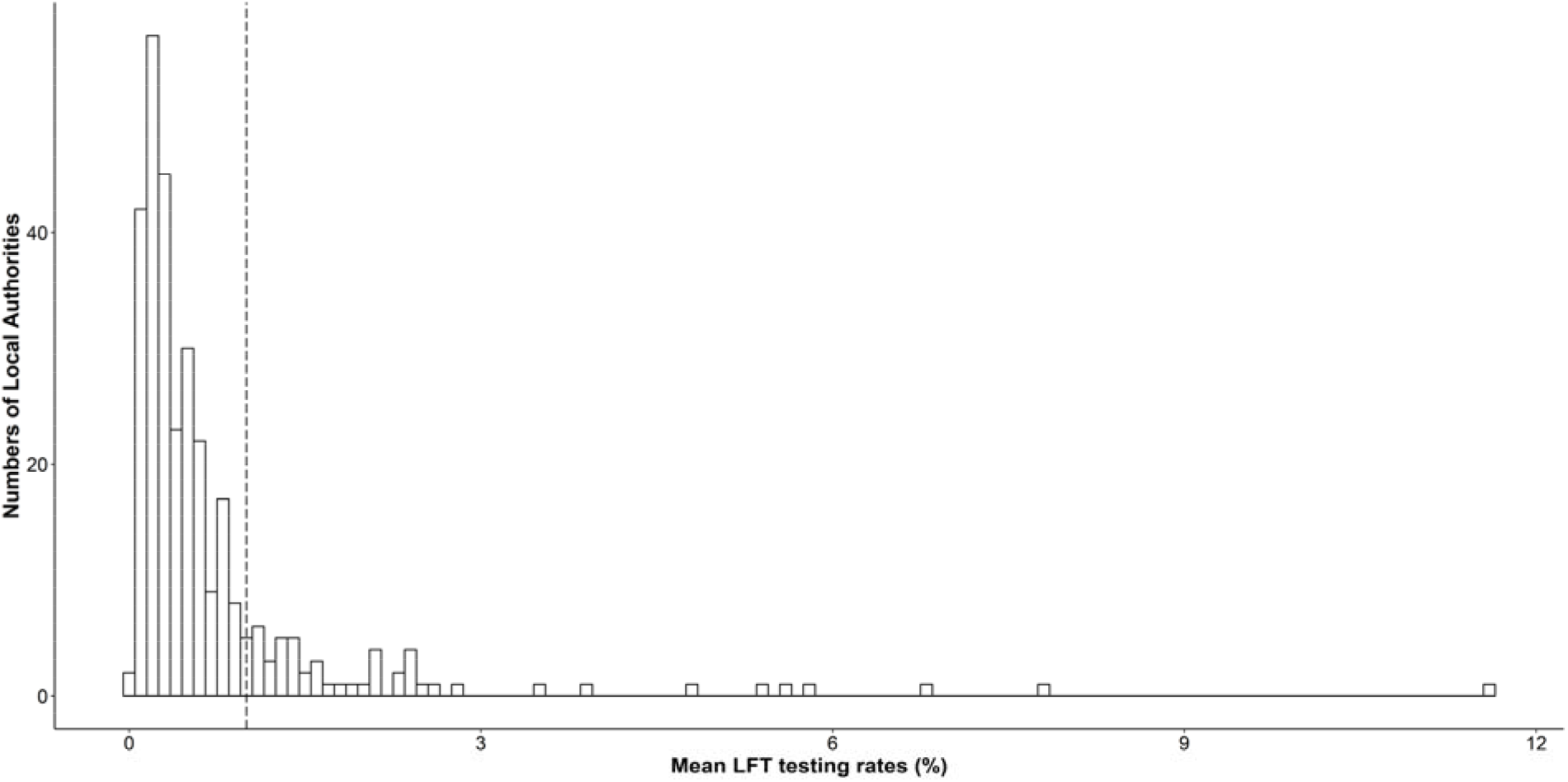
Distribution of mean weekly LFT tests per 100 population across Local Authorities in England between 6^th^ November 2020 and 2^nd^ January 2021. Note: Dotted vertical line identifies the threshold of the mean LFT testing rate of 1 per 100 population per week that we used to exclude MSOAs from the control group to minimise the potential impact of similar asymptomatic pilot programmes elsewhere.

### Part 2: Estimating Tier 3 restrictions on SARS-CoV-2 transmission compared to entering Tier 2 restrictions on 3 December 2020

We analysed the impact of Tier 3 restrictions on SARS-CoV-2 transmission introduced in December 2020 in England, compared to Tier 2 restrictions. The main differences of Tier 2 and 3 were additional restrictions on meeting people outdoors and restrictions on the hospitality sector. To be more specific, in Tier 3 people were prohibited from meeting with people outside their household in private gardens whilst in Tier 2 people were allowed to meet with up to six people in private gardens; pubs and restaurants were closed in Tier 3 areas whilst in Tier 2 areas only those serving food remained open. We used data on COVID-19 restrictions that were compiled and made available by the Open Data Institute.[32]

As Liverpool introduced the less restrictive Tier 2 measures on 3^rd^ December 2020, whilst new tier’s restrictions typically took effect in many local authorities on the first forthcoming Monday (7^th^ December 2020), we therefore specified 2 weeks after that point as the earliest plausible time at which the change in restrictions could affect hospital admissions (20^th^ December 2020). We investigated the change in MSOA-level COVID-19 hospital admissions in the intervention group (Tier 3 areas) using synthetic control analysis, 12 weeks before and 10 weeks after that time point, compared to a synthetic control group derived from places that entered Tier 2 at the same time. The exact time frame ranged from 4 October 2020 to 21 February 2021. We identified 2,809 Tier 3 MSOAs as the intervention group, whilst the synthetic control group was constructed from the 3,481 Tier 2 MSOAs (excluding 61 MSOAs in the Local Authority of Liverpool).

Along with the local area characteristics outlined above in our analysis of SMART, we additionally accounted for differences in the prevalence of the new variant B.1.1.7, which became dominant during that time period, by including the proportion of positive tests with S-gene failure on PCR testing for each local authority in the period before the 17^th^ December using data from Public Health England.[23] To further control for the confounding effect of community testing, we also excluded the 142 MSOAs (2.2% of all non-intervention MSOAs) from the control group if they were within authorities with a mean LFT testing rate of more than 1 per 100 population per week between 6th November and 2nd January 2021. In the sensitivity testing in Part 5 of SF, we provided the estimation of the Tier 3 effect upon hospitalisation, without excluding these 142 MSOAs.

The introduction of Tier 3 restrictions in December was associated with an average reduction of hospital admissions of 17% (95% CI 13% to 21%) in Tier 3 areas compared to Tier 2 areas. In the sensitivity testing in Part 5 of SF, without excluding the 142 MSOAs with a mean LFT testing rate of more than 1 per 100 population per week during the pilot period of the community testing in Liverpool, the introduction of Tier 3 restrictions in December was associated with an average reduction of hospital admissions of 14% (95% CI 9% to 17%) in Tier 3 areas compared to Tier 2 areas.

As can be seen from Figure SF3 the effect of the Tier restrictions changes over time. In adjusting the MSOA hospital admissions in Tier 3 areas to account for this in our analysis of SMART, we applied the weekly effect sizes from the synthetic control analysis of Tiered restrictions.

**Figure SF3.**
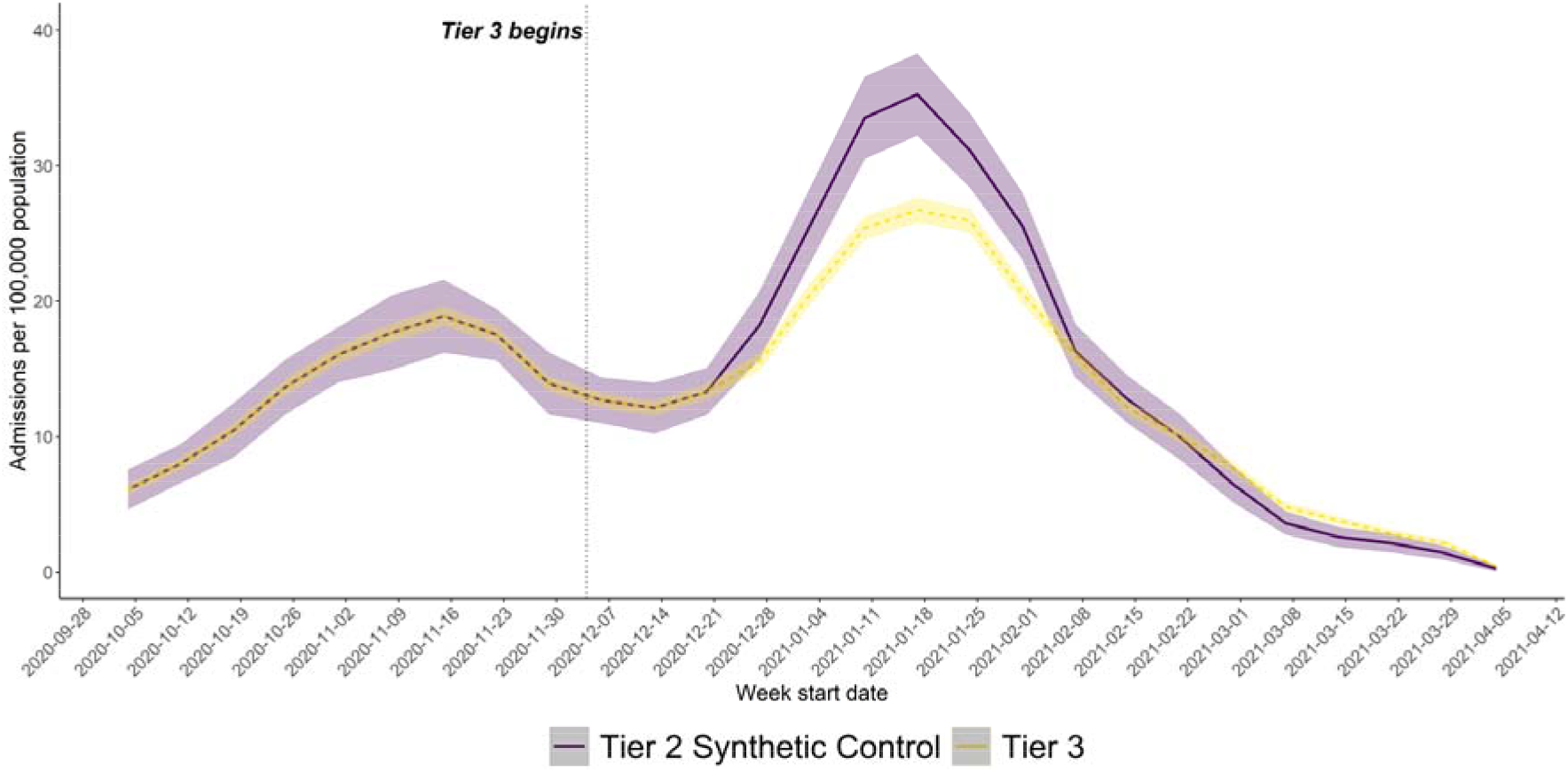
Chart showing the effect of Tier 3 restrictions on COVID-19 hospital admissions relative to the Tier 2 synthetic control group.

### Part 3: Synthetic control area construct weighting

Figure SF4 shows the geographical pattern of MSOAs used to construct the synthetic control group. The intervention group (61 MSOAs in Liverpool) is coloured in black, whilst the synthetic control group is constructed by applying the calibrated weights to various MSOAs across the country. Most of these MSOAs constituting the synthetic control group are in the North West region and along the North East coastlines.

**Figure SF4.**
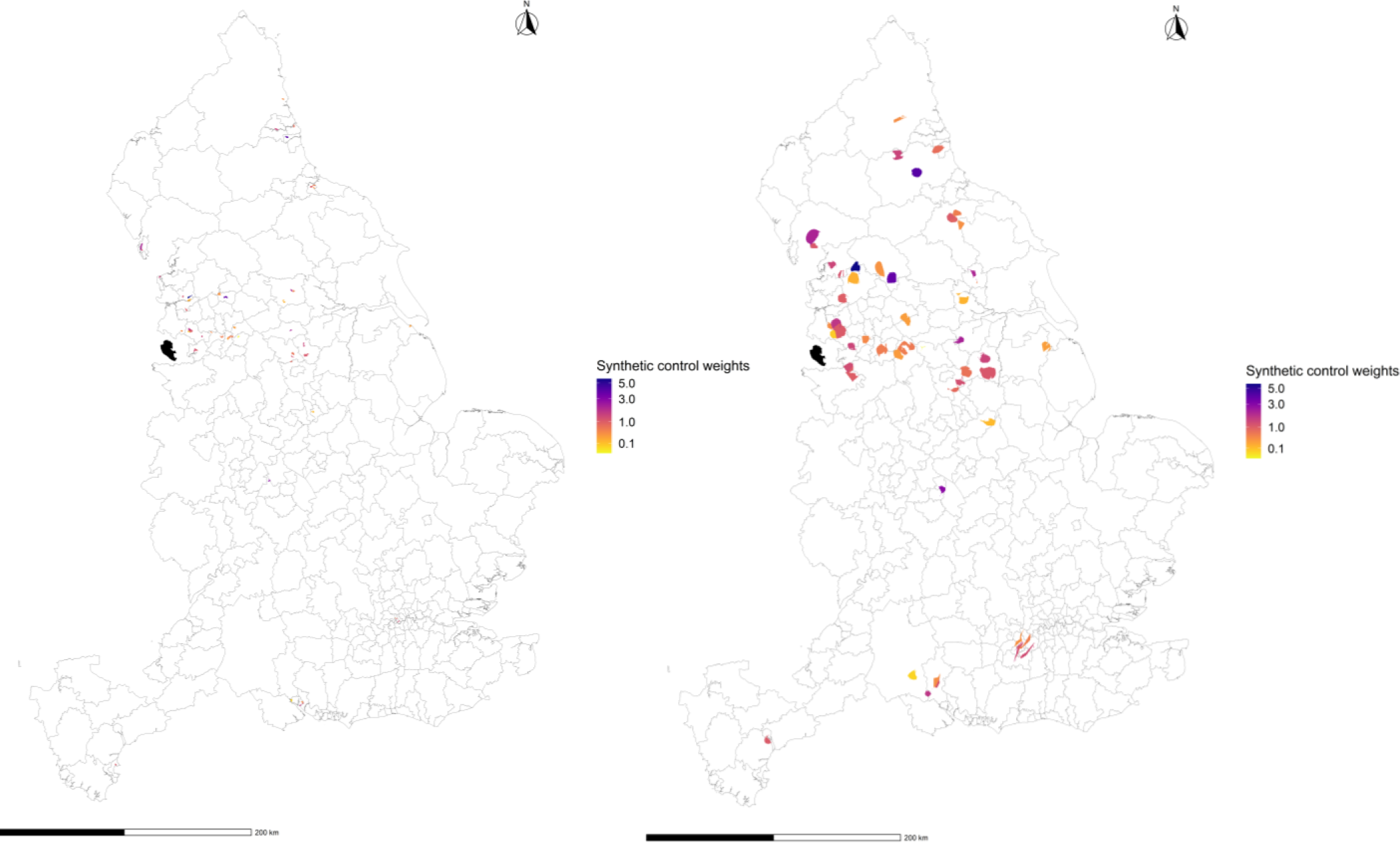
Weighting of areas used to construct the synthetic control group (Liverpool, coloured in black). The maps relate to a particular seed draw, with 54 MSOAs contributing: the left map shows the synthetic control MSOAs in their original locations and sizes; the right map enlarges these MSOAs proportionally to their synthetic control weights (hence inaccurate locations) to facilitate understanding.

**Table SF1.** Estimated effect of community testing programme on COVID-19 hospital admissions from synthetic control analysis, adjusting the Tier 3 effect by the 95% upper and lower bound during the pilot period. This estimation is made under alternative assumptions related to the effect of the introduction of less stringent Tier 2 restriction in Liverpool in December 2021, excluding the 142 MSOAs with an LFT testing rate of more than 1 per 100 population per week during the pilot period.

| **Model** | **Intervention period** | **Assumed reduction in COVID-19 hospital admissions from Tier 3 vs 2 restrictions** | **% difference in COVID-19 hospital admissions between Liverpool and control** | **Lower 95% CL** | **Upper 95% CL** | **P-value** |
| --- | --- | --- | --- | --- | --- | --- |
| **3A** | 6<sup>th</sup> November 2020 to 2<sup>nd</sup> January 2021 | 21% (upper 95% bound) | - 29% | - 39% | - 15% | <0.001 |
| **3B** |  | 13% (lower 95% bound) | - 22% | - 31% | - 7% | 0.014 |

### Part 5: Sensitivity analysis: excluding controls with high lateral flow testing

**Table SF2.** Estimated effect of community testing programme on COVID-19 hospital admissions from synthetic control analysis, without excluding MSOAs with LFT testing rates of more than 1 per 100 population per week during the pilot period from the control group. This estimation is made under alternative assumptions related to the effect of the introduction of less stringent Tier 2 restriction in Liverpool in December 2021, including the 142 MSOAs with an LFT testing rate of more than 1 per 100 population per week during the pilot period.

| **Model** | **Intervention period** | **Assumed reduction in COVID-19 hospital admissions from Tier 3 vs 2 restrictions** | **% difference in COVID-19 hospital admissions between Liverpool and control** | **Lower 95% CL** | **Upper 95% CL** | **P-value** |
| --- | --- | --- | --- | --- | --- | --- |
| **A** | 6<sup>th</sup> November to 3<sup>rd</sup> December 2020 | Nil | - 45% | - 54% | - 34% | <0.001 |
| **B** | 6<sup>th</sup> November 2020 to 2<sup>nd</sup> January 2021 | Nil | - 11% | - 22% | 5% | 0.182 |
| **C** |  | 14% (central estimate) | - 20% | - 29% | - 6% | <0.001 |
| **D** |  | 17% (upper 95% bound) | - 24% | - 33% | - 11% | <0.001 |
| **E** |  | 9% (lower 95% bound) | - 17% | - 26% | - 2% | 0.040 |

## Notes

### Author Declarations

All the data we used are publicly accessible from https://coronavirus.data.gov.uk/.

